# An Overview of Retraction Status and Reasons of Non-Cochrane Systematic Reviews in Medicine

**DOI:** 10.1101/2020.10.10.20210666

**Authors:** Qianling Shi, Zijun Wang, Qi Zhou, Ruizhen Hou, Xia Gao, Shaoe He, Siya Zhao, Yanfang Ma, Xianzhuo Zhang, Quanlin Guan, Yaolong Chen, on behalf of PRESS working group

**Author notes:** **Correspondence at:** Yaolong Chen. Evidence-Based Medicine Center, School of Basic Medical Sciences, Lanzhou University, Lanzhou 730000, China.; Quanlin Guan. Department of Oncology Surgery, The First Hospital of Lanzhou University, Lanzhou 730000, China.

## Abstract

**Background:** Many previous studies have analyzed the status of retracted publications from different perspectives, but so far no study has focused on systematic reviews (SRs). The purpose of this study is to analyze the retraction status and reasons of non-Cochrane SRs in the field of medicine.

**Methods:** We searched MEDLINE and Embase from their inception to April 18, 2020, as well as Retraction Watch Database and Google Scholar with no language restriction to find non-Cochrane SRs that were retracted for any reason. Two reviewers independently screened and extracted data. We describe the characteristic and reasons of retraction and the duration from publication to retraction.

**Results:** We identified 150 non-Cochrane SRs in medicine retracted between 2004 and 2020. The majority of retracted SRs were led by authors from China and affiliated with hospitals. Most SRs were published in journals with an impact factor ≤3, and in journal ranked in the third quarter. The largest proportion of retraction notices were issued by the publisher and editor(s) jointly; seven did not report this information. Fraudulent peer-review was the most common reason for retraction, followed by unreliable data meaning errors in study selection or data analysis. The median time between publication and retraction was 14.0 months. SRs retracted due to research misconduct took longer to retract than SRs retracted because of honest error.

**Conclusions:** The situation with retracted SRs is critical globally, and in particular in China. The most common reasons for retraction are fraudulent peer-review and unreliable data, and in most cases the study is retracted more than a year after publication. Efforts should be made to improve the process of peer review and adherence to the COPE retraction guidance, while at the same time authors should strengthen their skills in SR methodology.

## Background

Retractions of published articles are sometimes needed to maintain the integrity of the scientific literature and to alert the reader of potential serious problems identified within the article ^[1,2]^. The Committee on Publication Ethics (COPE) published retraction guidance in 2009. Reasons for retraction include unreliable data, redundant publication, plagiarism, and failure to disclose conflict of interest. Unreliable data may result from honest error (such as mis-calculation or experimental error) or from research misconduct (such as data fabrication) ^[2]^. A study published in 2011 found that in the past decade, the number of retraction notices has shot up 10-fold, even as the literature has expanded by only 44% ^[3]^. Another study published in 2019 showed that of the 21,859,178 publications indexed in the Web of science between 1978 and 2017, 2,859 were retracted, of which, the United States has the most retractions, but China has the highest retraction rate ^[4]^. Retractions involved papers led by researchers from over 50 countries worldwide ^[5]^. Having a paper retracted does not only affect the academic status and reputation of individual authors, leading possibly to administrative, civil, and criminal sanctions, but also correlates with decreased productivity, funding and resources ^[5-7]^. Retractions of publications in the field of biomedicine also propose a great threat to public health ^[8]^.

Systematic Reviews (SR), which are being widely used in the domain of medicine, have had a revolutionary impact in many fields of science, helping to establish evidence-based practice ^[9,10]^. Scientifically rigorous SRs, carried out following formal protocols, have long been considered at the top of the hierarchy of evidence-based medicine studies. SRs integrate information, address well-defined questions and lead to new insights, but also provide a robust overview of a problem or field of research, and serve as important evidence for development of clinical practice guidelines. Therefore, SRs are an essential part of clinical decision-making and research ^[11,12]^. To avoid waste of research, no new studies should be done without a SR of existing evidence ^[13]^. However, while the number of published and registered SRs has increased substantially over the past 20 years, the extent to which their implications are correctly understood is poor. The majority of produced SRs are unnecessary, misleading, or conflicted ^[14-18]^, and this is a particular problem among non-Cochrane SRs used less rigorous methods than Cochrane reviews ^[19]^. In recent years, there is a growing call to reduce the number of articles that need to be later retracted.

To the best of our knowledge, no research has yet specifically aimed to describe the main features of retracted SRs. Therefore, on behalf of PRESS (Publication Science of Retracted Studies) working group, present study aims to perform an in-depth analysis of the status and reasons of retracted non-Cochrane SRs and meta-analyses in the field of medicine.

## Methods

### Search strategy

Two reviewers (Qianling Shi and Zijun Wang) comprehensively searched MEDLINE (via PubMed) and Embase from their inception through April 18th, 2020. No language restriction was applied. We conducted the search by combining the MeSH and free words including “Retraction”, “Withdraw”, “Remove”, “Meta-analysis”, “Systematic Review” and their derivatives. The search strategy was also peer reviewed by an external specialist. Details of the search can be found in the ***Supplementary Material I***. We also retrieved data from the Retraction Watch Database (RWdb, www.retractionwatch.com) and Google Scholar (https://scholar.google.nl/) for relevant studies that met the inclusion criteria. RWdb is a public platform for retractions of scientific papers, it is considered as the largest and most comprehensive database of retracted articles ^[20]^.

### Inclusion and exclusion criteria

We included all non-Cochrane SRs published in the field of medicine that were retracted. The corresponding retraction notices were also extracted for analysis. Studies were excluded if they were: 1) duplicates; 2) use of the words “retraction” or “retracted” but with a different meaning from the one considered by the present review; 3) topic unrelated to the field of medicine; 4) topic related to the field of medicine in animals; 5) retracted Cochrane SRs; and 6) retracted SRs that were republished.

### Study selection

Two reviewers (Qianling Shi and Zijun Wang) independently screened first the titles and abstracts after eliminating duplicates, and then the full-texts for potentially relevant articles, using pre-defined criteria. The specific bibliographic software EndNote X9 was used. Discrepancies were discussed, if necessary with a third researcher (Qi Zhou). The process of study selection was documented using a flow diagram.

### Data extraction

Two reviewers (Qianling Shi and Zijun Wang) independently extracted the following data with a standardized data collection form: 1) basic information (title of retracted article, country and affiliation of the first author, year and month of online publication and retraction); 2) journal information (journal name, impact factor [IF] of 2018 according to ISI Web of Knowledge, quartile in Journal Citations Reports (JCR) of Thomson Reuters of 2018); 3) retraction characteristics (who retracted the article [the decision to retract a paper can be made by editors, authors or the author’s employer, publisher or others], reasons for retraction, availability of retraction notice).

After considering classifications of retractions in previous studies, retraction reasons for SRs were classified into the following categories, and definitions are as follows ^[1,2,5,21]^.

1. Repetitive research: the content of the retracted article is the same as that of a previously published paper;
2. Data falsification: data has been manipulated or made up;
3. Duplicate publication: the paper had been published more than once or there was significant overlap between manuscripts (usually as a result of author misconduct);
4. Authorship issues: not all authors aware of manuscript submission;
5. Plagiarism: duplication of text from previously published articles;
6. Unreliable data: data has error due to miscalculation or experimental error; in the case of SRs, this can be an error in study selection, data collection, data analysis, result interpretation, etc.
7. Fraudulent peer-review: it can arise when editors rely on authors’ recommended reviewers. The names are often genuine but have a false e-mail address that enables the authors to write a favourable review of their own paper, thereby facilitating acceptance.
8. Others: reasons that cannot be divided into the above categories.

All reasons were classified by one reviewer (Qianling Shi) and checked for agreement by another reviewer (Zijun Wang) using the information given in the retraction notice, any discrepancies were settled by discussion. If we were unable to retrieve the notice, we searched the RWdb to find information about the reasons and characteristics of retraction. When multiple reasons for the retraction were given, all were described and noted.

### Statistical analysis

Descriptive statistics of the retraction characteristic, reasons for retraction, and time to retraction (defined as the time from original publication date to retraction date and) of articles were calculated. Two independent reviewers (Qianling Shi and Zijun Wang) analysed all datasets with SPSS 18 Software. Comparisons of the causes for retraction among sub-groups (IF, journal rank in JCR) were performed using the Chi square test or variance analysis, and t test was used to compare the time from publication to retraction. Statistical significance was set at *P* < 0.05. Categorical data were reported using frequencies and/or percentages. Continuous variables were described using median with range (minimum to maximum) or interquartile range (IQR). We also analyzed the adherence to COPE guidance on retraction in all respects (which suggest that retractions be linked to the retracted article, be clearly identified as a retraction, published promptly and freely available to all readers. Retractions should also state who is retracting the article, reason(s) for retraction and avoid statements that are potentially defamatory or libellous, of which, issuers can be author(s), journal editor or journal’s owner such as a learned society or publisher) ^[2]^.

## Results

### Search results

Electronic and additional search resulted in a total of 1,945 records (1,453 in MEDLINE, 430 in Embase and 62 in RWdb). Of these records, 945 were excluded as duplicates. After screening the titles, abstract and full texts, a total of 150 non-Cochrane SRs in the field of medicine retracted between 2004 and 2020 were included. The process of study selection including the reasons for exclusion is documented in Figure 1.

**Figure 1.**
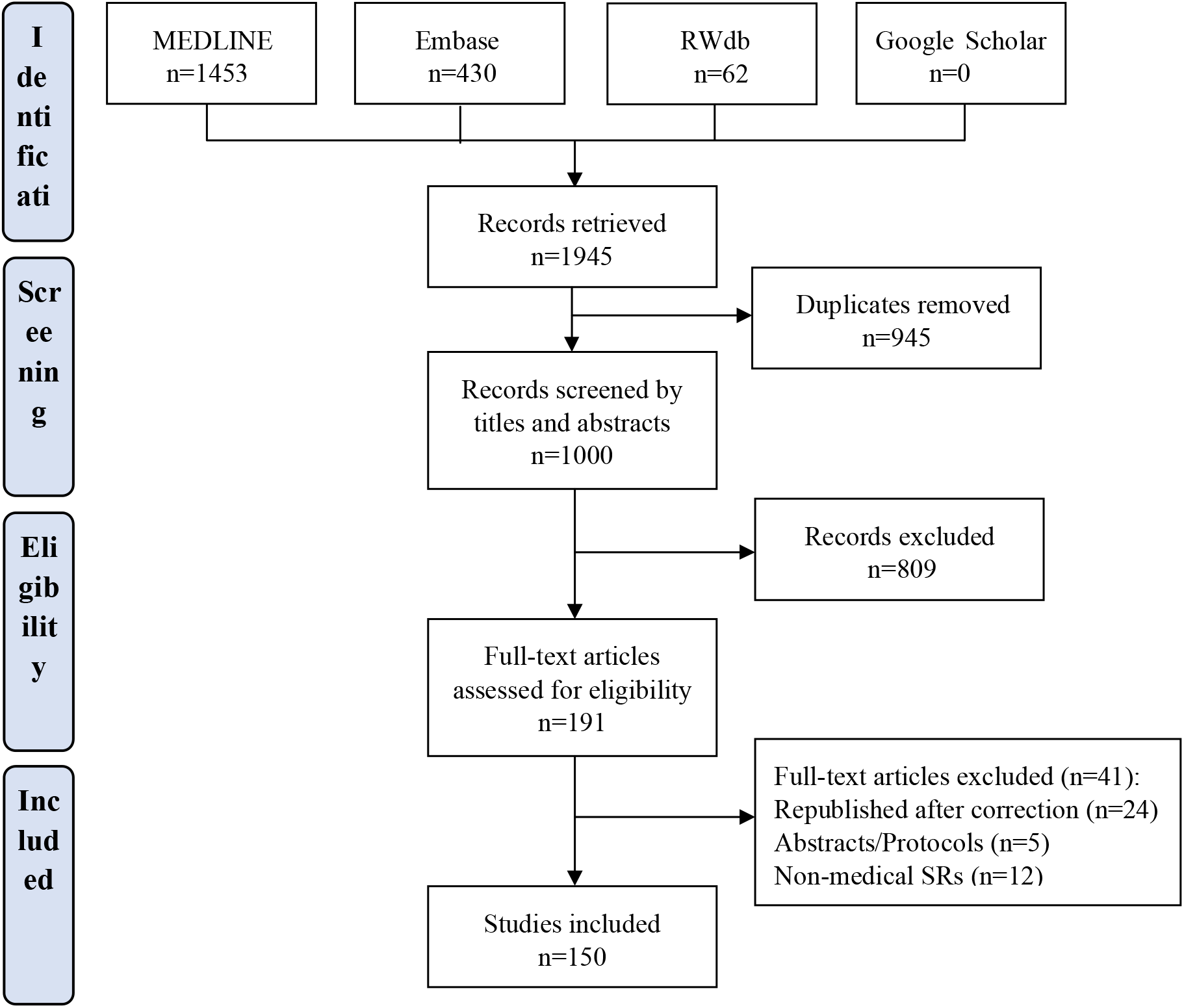
Flow diagram of the literature search.

### Study characteristic

The characteristics of the 150 included studies are displayed in Table 1. As shown in Figure 2, with the total number of published SRs is large and growing over the past decade (from 2011 to 2020), the total number of retracted SRs is small but increased as well. The first SR, published in 2003, was retracted in 2004, and the highest retraction rate was in 2015 (n=43, 28.7%). The first authors of retracted SRs were from 18 countries, most from Asia (n=126, 84.0%). China was the leading country of origin for the total number of retracted publications (n=113, 75.3%), followed by the United States (n=5, 3.3%), Italy (n=5, 3.3%), the United Kingdom (n=4, 2.7%) and Iran (n=4, 2.7%). The most frequent type of organization of the first author was hospital (n=106, 70.7%), followed by university (n=41, 27.3%).

**Table 1.** Characteristic of retracted systematic reviews (N=150)

| Categories | Number (Percentage) |
| --- | --- |
| **Year of retraction** |  |
| 2015~2020 | 137 (91.3%) |
| 2010~2014 | 11 (7.3%) |
| 2005~2009 | 1 (0.7%) |
| 2003~2004 | 1 (0.7%) |
| **Country of first author** |  |
| China | 113 (75.3%) |
| USA | 5 (3.3%) |
| Italy | 5 (3.3%) |
| UK | 4 (2.7%) |
| Iran | 4 (2.7%) |
| Japan | 3 (2.0%) |
| Brazil | 3 (2.0%) |
| Korea | 2 (1.3%) |
| Singapore | 2 (1.3%) |
| Others* | 10 (6.7%) |
| **Organization** |  |
| Hospital | 106 (70.7%) |
| University | 41 (27.3%) |
| Others | 3 (2.0%) |
| **Journal†** |  |
| *Tumor Biol* | 23 (15.3%) |
| *Mol Biol Rep* | 13 (8.7%) |
| *Medicine* | 9 (6.0%) |
| *Mol Neurobiol* | 9 (6.0%) |
| *Plos One* | 6 (4.0%) |
| *Eur J Med Res* | 5 (3.3%) |
| *J Orthop Surg Res* | 5 (3.3%) |
| *Others* | 80 (53.3%) |
| **Impact Factor (IF) of journal** |  |
| 0 < IF ≤ 3 | 85 (56.7%) |
| 3 < IF ≤ 5 | 22 (14.7%) |
| IF > 5 | 13 (8.7%) |
| Not indexed in SCI‡ | 30 (20.0%) |
| **Quartile in Journal Citation Report** |  |
| Q1 | 32 (21.3%) |
| Q2 | 35 (23.3%) |
| Q3 | 39 (26.0%) |
| Q4 | 14 (9.3%) |
| Not indexed in SCI‡ | 30 (20.0%) |
| **Remarked as retracted article** |  |
| Yes | 148 (98.7%) |
| No | 2 (1.3%) |
| **Retraction notice** |  |
| Published | 148 (98.7%) |
| Unpublished | 2 (1.3%) |

**Note** \* Others included Canada (n=1), Colombia (n=1), New Zealand (n=1), Sweden (n=1), Denmark (n=1), Malaysia (n=1), Argentina (n=1), Saudi Arabia (n=1), Nigeria (n=1),
† Journal was presented in their abbreviations
‡ SCI: Science Citation Index

**Figure 2.**
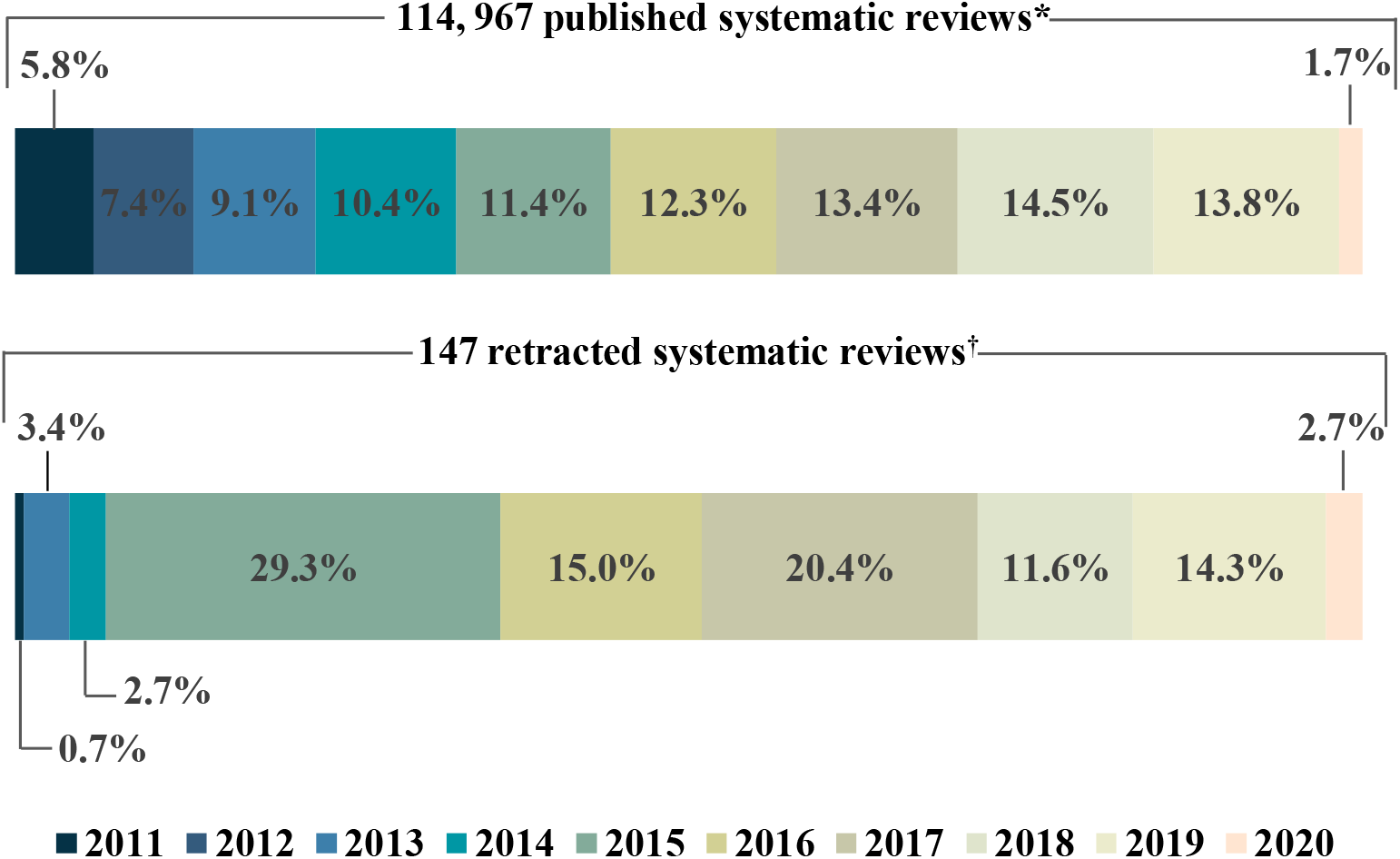
Proportion of published and retracted publications in relation to the total annual number systematic reviews over the past decade. *Data for published studies related to systematic reviews retrieved on 18 April 2020 by searching for non-Cochrane systematic reviews in the field of medicine in the MEDLINE. ^**†**^Smaller than total number of retractions because it only included retracted systematic reviews between 2011 to 2020.

A total of 79 journals were involved. *Tumor Biology* retracted the most (n=23, 15.3%), followed by *Molecular Biology Reports* (n=13, 8.7%), *Medicine* (n=9, 6.0%) and *Molecular Neurobiology* (n=9, 6.0%). All included SRs were written in English (except one that was published in Chinese. Most articles (n=120, 80.0%) were published in journals listed in the Science Citation Index (SCI). Most retracted SRs (n=85, 56.7%) were published in low impact (≤3) journals, and most were from journal ranked in the third highest quarter (n=39, 26.0%), followed by journals in the second highest quarter (n=35, 23.3%).

Not all retraction notices were clearly labelled and linked to the retracted articles. For a total of 148 (98.7%) studies a retraction notice was published, of which the first in 2004. For two (1.3%) studies no retraction notice was issued, but the study was presented with a watermark to identify the article was retracted. For these studies, we determined the issuer of the retraction notice as unclear.

### Retraction characteristic

#### Issuer of the retraction notice

We categorized the entities that retracted the 150 SRs into nine groups (Table 2). The largest proportion of retraction notices were issued by the publisher and editor(s) jointly (n=60, 40.0%), followed by the editor alone (n=27, 18.0%) and author alone (n=16, 10.7%). For nine (6.0%) included studies, we were unable to determine who issued the retraction notice. Of the eight notices, two were from the author(s) and/or editor, one was from publisher or journal, three used the words as “we”, one used the words as “they”, and for two no notice was issued. Although most retraction notices declared who retracted the article, 7 (4.7%) did not explicitly state this information.

**Table 2.**
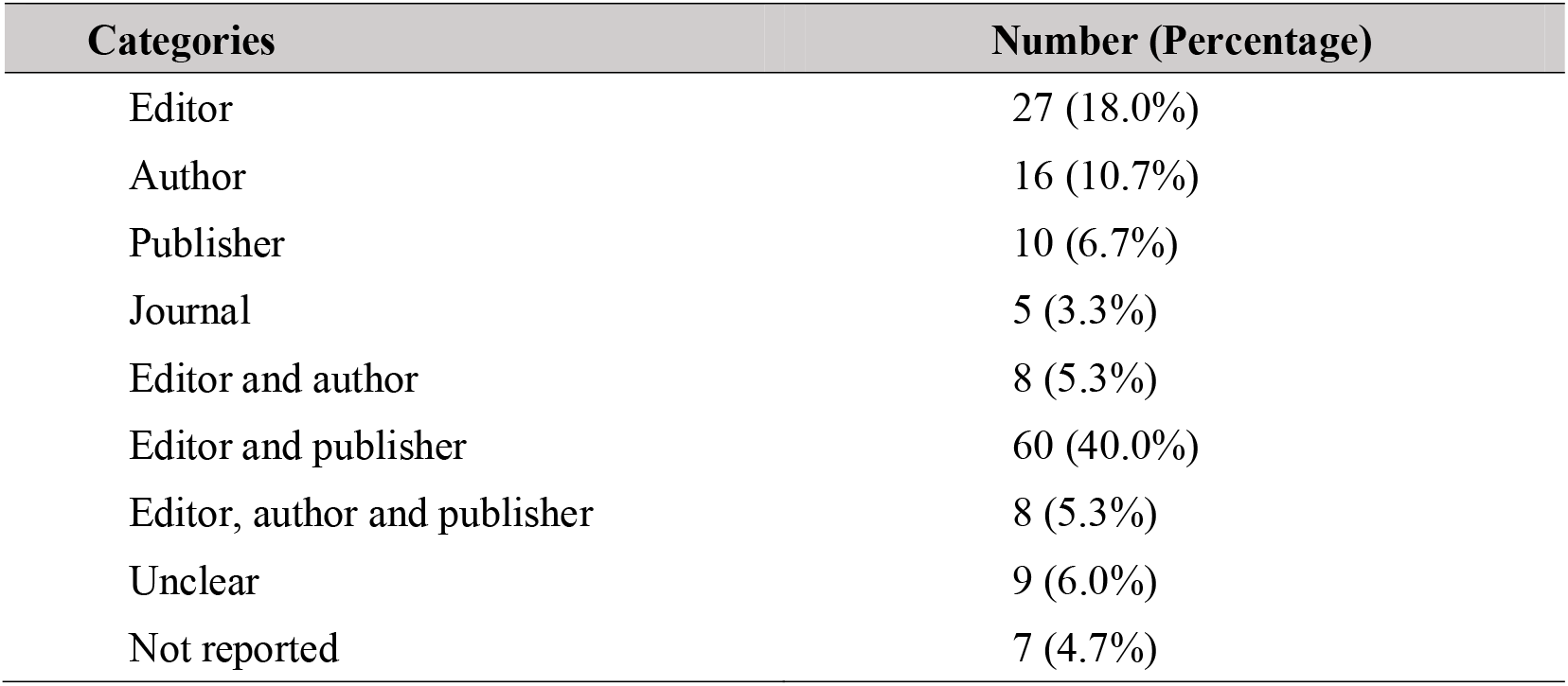
Number of retractions listed by the issuer (N=150)

#### Reasons of retraction

A total of 161 reasons for retraction were mentioned for the included 150 SRs (Table 3). We divided the retraction reasons into three categories: honest error (unreliable data), misconduct (data fabrication, plagiarism, duplicate publication, authorship issues, compromised peer review), and unclear (not possible to distinguish “honest error” from “misconduct”). The vast majority of retracted SRs (n=100, 66.7%) were due to some form of research misconduct.

**Table 3.**
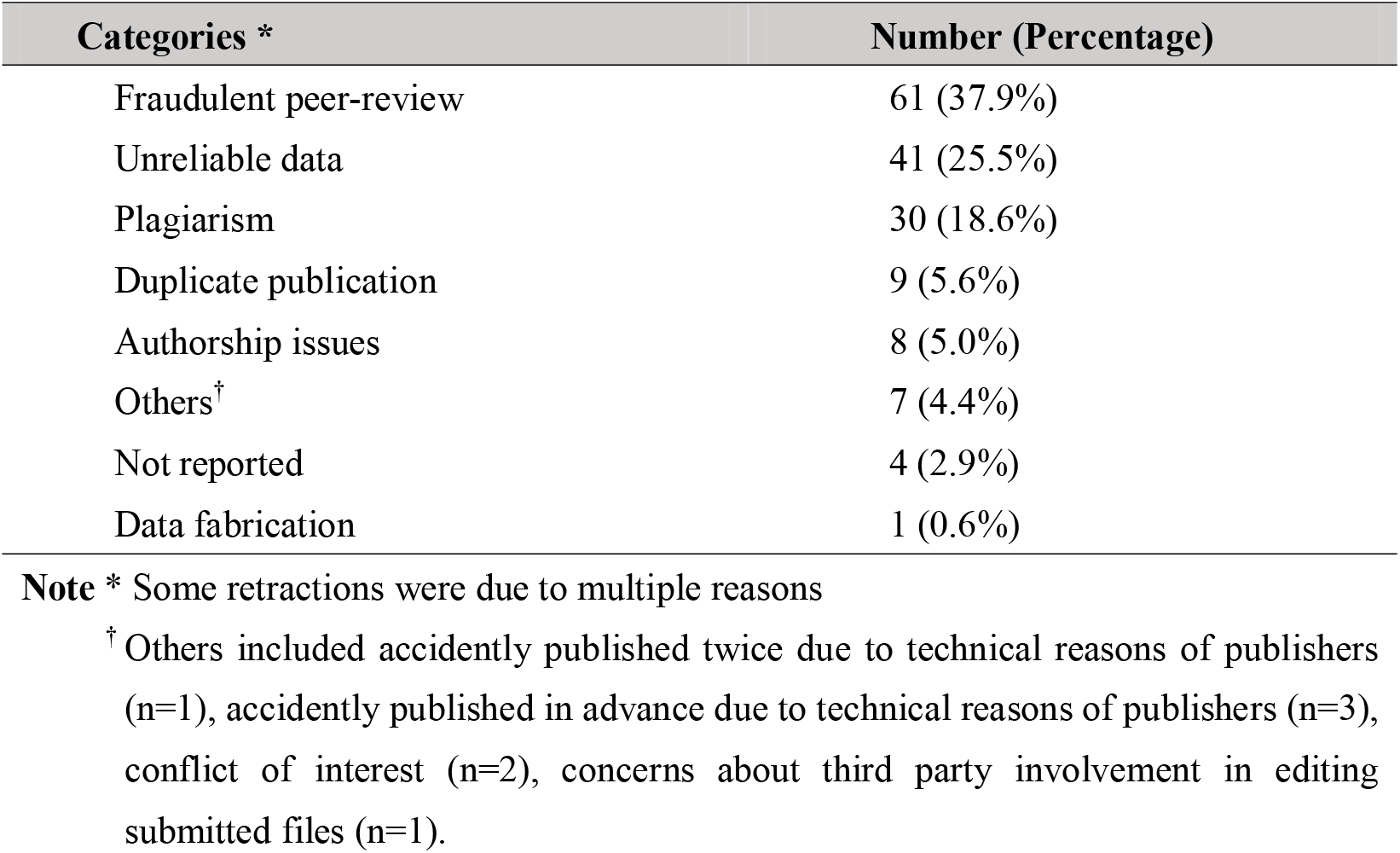
Reasons of retraction (N=161)

The most common reason for retraction was fraudulent peer-review (n=61, 37.9%), followed by unreliable data (n=41, 25.5%) and plagiarism (n=30, 18.6%). Most reasons classified as unreliable data were due to error in either the selection of included studies, or in data analysis (Table 4). Other reasons included duplicate publication (n=9, 5.6%), authorship issues (n=8, 5.0%) and only one (0.6%) was retracted due to data fabrication. No articles were retracted due to repetitive research. For some retraction statements, which appeared to use deliberately ambiguous wording that made it difficult to distinguish honest errors from suspected (or proven) misconduct, we assigned into others (n=7, 4.4%). No reason was provided for four (2.5%) retractions. Figure 3 shows the growth and variation in reasons for retractions over time.

**Table 4.** Detailed reasons of retraction due to unreliable data.

| Categories * | Number |
| --- | --- |
| **Error in search strategy** | **4** |
| • Inadequate reporting of search strategy | 3 |
| • Lack of key search items | 1 |
| **Error in selection of included studies** | **18** |
| • Included studies that did not meet the inclusion criteria | 9 |
| • Included duplicate studies | 5 |
| • Included studies that were retracted | 3 |
| • Included unpublished studies accessible only during peer review for another journal | 1 |
| • Concerns in inclusion/exclusion criteria | 1 |
| • Concerns in reasons for excluded studies | 1 |
| **Error in data collection** | **10** |
| • Error in study design <sup>†</sup> | 3 |
| • SE were extracted instead of SD | 2 |
| • Data in some figures were not found in the primary article | 1 |
| • Exact reason not reported | 5 |
| **Error in data analysis** | **16** |
| • Incorrect methods of constructing forest plots | 7 |
| • Incorrectly labelled figures | 2 |
| • Incorrect analysis of publication bias | 2 |
| • Incorrect subgroup analyses | 1 |
| • Lack of sensitivity analyses | 1 |
| • Lack of adequate analyses for some outcomes | 2 |
| • Exact reason not reported | 6 |
| **Error in data reporting <sup>‡</sup>** | **2** |
| **Error in result interpretation** | **4** |
| • Incorrect conclusions | 3 |
| • Exact reason not reported | 1 |
| **Others** | **7** |
| • Multiple errors in the methodology | 5 |
| • Incorrect use of the reporting guidelines | 2 |
| • Errors in reference list | 2 |

**Note** : SE: Standard Error; SD: Standard Deviation.
\* Some retractions were due to multiple reasons.
† Error in study design included error data in experimental groups included in the study, the number of study arms in the trial, number of participants reported, treatment duration, etc.
‡ Data provided in the figure were not consistent with those shown in result or abstract sections.

**Figure 3.**
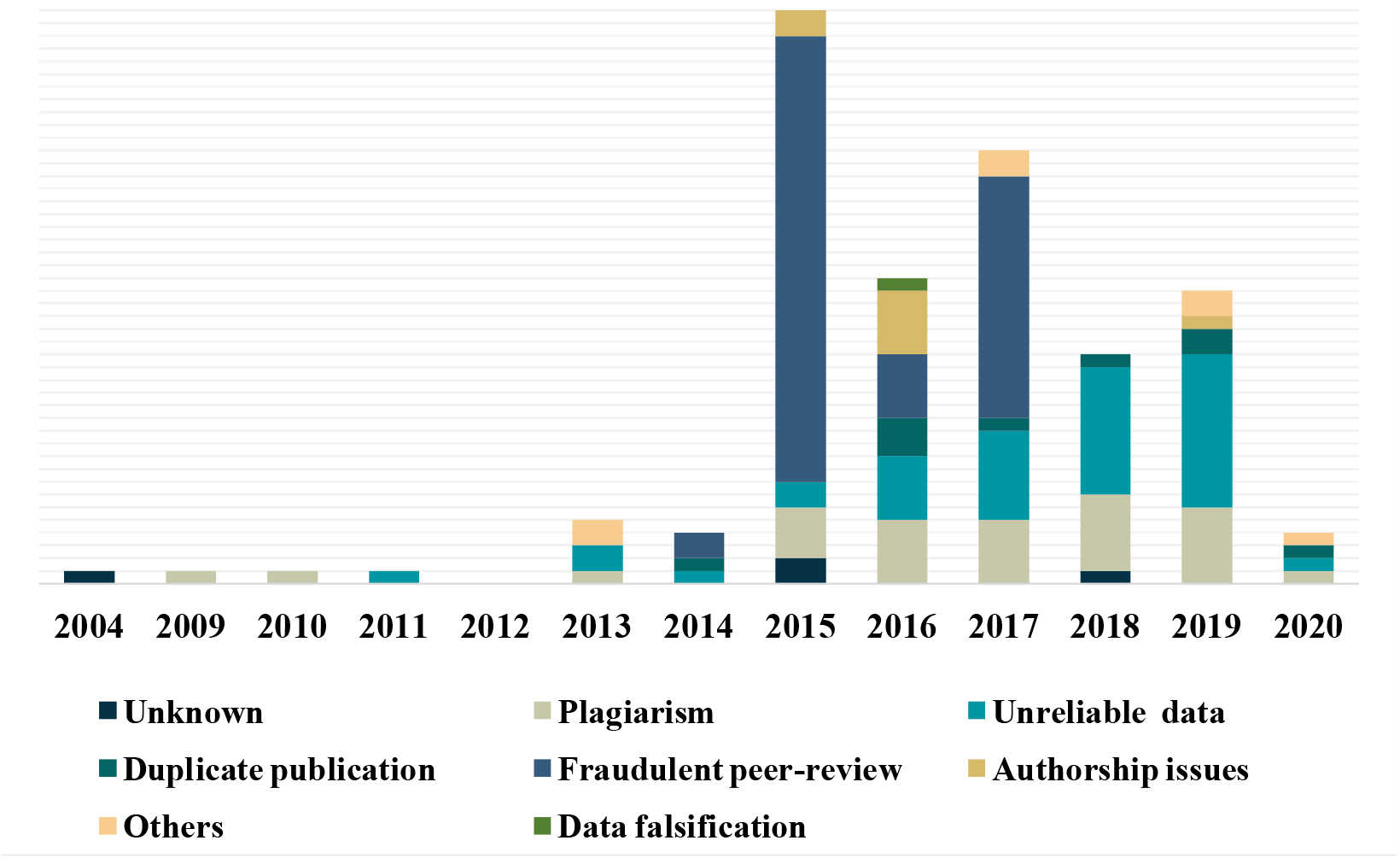
The annual retractions by reason from 2004 to 2020.

In 11 (7.3%) studies two reasons for retraction were given. Among the 11 retracted SRs, four were retracted due to fraudulent peer-review and plagiarism, two were retracted due to plagiarism and authorship issues, two were retracted due to fraudulent peer-review and authorship issues, one was retracted due to plagiarism and authorship issues, one due to plagiarism and unreliable data, and one due to unreliable data and concerns about third party involvement in editing submitted files.

In addition to this, causes for retraction also varied according to journal IF (*P*=0.000) and journal rank in JCR (*P*=0.000): retractions due to unreliable data was most frequent from journals with a high IF (equal or more than 5) or high journal rank (Q1 and Q2), while a faked review process was the main cause of retraction from journals with a low IF (less than 5), low journal rank (Q3 and Q4) and journals with no citation index. In-depth analysis showed that the most common reason for retraction in hospital and university was fraudulent peer-review and unreliable data, respectively.

#### Retraction interval

The median (range) time between publication of the 150 SRs and their retraction was 14.0 (0.0∼108.0) months, of which, most of the included SRs (n=105, 70.0%) were retracted within 20 months after publication. Seven SRs were retracted within one month, of which, five on the same day the article was published, we therefore determined the month to retraction was zero.

The duration from publication to retraction also varied by the reasons of retraction (Table 5). When excluding the reason of data fabrication (only one study with an interval of 75 months), authorship issues resulted the longest with a minimum of six month and a maximum of 50 months, followed by fraudulent peer-review, plagiarism, duplicate publication and unreliable data, The median (IQR) months of retraction interval for fraudulent peer-review was longer in studies published before 2015 (18.0 [13.0∼37.5]) than after 2015 (16.0 [9.5∼16.0]), but not significantly with *P*=0.249. In general, SRs with research misconduct (including data fabrication, fraudulent peer-review, authorship issues, plagiarism, and duplicate publication) took longer to be retracted (16.0 [10.00∼31.3] months) than honest error (7.0 [4.0∼22.0] months) in total, *P*=0.000.

**Table 5.** Time from publication to retraction by cause for retraction.

| Categories* | Months to retraction<sup>†</sup> |
| --- | --- |
| Authorship issues | 27.5 (6.0~50.0) |
| Fraudulent peer-review | 16.0 (2.0~51.0) |
| Plagiarism | 15.5 (0.0~51.0) |
| Unreliable data | 7.0 (1.0~108.0) |
| Duplicate publication | 14.0 (0.0~40.0) |
| Data fabrication | 75‡ |
| Others | 3.0 (0.0~53.0) |
| Not reported | 2.5 (0.0~6.0) |
| Total | 14.0 (0.0~108.0) |

Note \* Some retractions were due to multiple reasons
<sup>†</sup> Data was showed by median (from minimum to maximum)
‡ Only one SR was retracted due to data fabrication

#### Reliability of retracted SRs

Retracted SRs have the potential to mislead current practice and future research through citations that occur before the publication of the retraction note, and erroneous citations that are made after the retraction. It is therefore important for researchers and readers to understand how much influence retracted SR can have on other studies. Our research group divided the reliability of retracted SRs into three categories:

1. High risk: The findings or conclusions of retracted SRs will be changed due to retraction so that citations will be severely affected (including unreliable data, data falsification).
2. Low risk: The findings or conclusions of retracted SRs will not be changed due to retractions so that citations will not be affected (including repetitive research, duplicate publication and authorship issues).
3. Unclear: we are uncertainty about whether the findings or conclusions of retracted SRs will be changed or whether citations will be affected (including compromised peer review and plagiarism)

The results in our study showed that 42 (28.0%) retracted SRs were considered as “high risk”, and only 12 (8.0%) as “low risk”.

## Discussion

Our study identified a total of 150 retracted non-Cochrane SRs in the field of medicine by 18 April 2020. The number of retractions steadily increased starting in 2004. The most common reasons were fraudulent peer-review and unreliable data. More than 75% were from China. Detailed analysis of the retraction notices revealed that COPE guidelines on retraction were not adhered to in all respects. In general, the mean number of months from publication to retraction was more than one year, and articles involving apparent misconduct took longer to retract than honest error.

The results from our study suggested that faked peer review was the most common reason for retraction. The number of retracted SRs reached the highest in 2015, it may be related to the large-scale retractions of academia happened in this year. The publisher *Springer Nature* retracted 64 articles in ten of its journals on the basis of fake peer review in August 18, 2015 ^[22]^. The retractions came only months after *BioMed Central* retracted 43 articles on similar grounds ^[23]^. Two years later, *Springer* retracted another 107 papers published in *Tumor Biology* ^[24]^. Data from the wake of large-scale retraction scandals and recent studies in other fields showed that most of the retractions were from China, and majority of SRs were from hospitals and published in journals with low IF. *Tumor Biology* and *Molecular Biology Reports* retracted the largest number of studies, the results were similar to those identified in other studies ^[5,22-24, 25-29]^. SRs involving research misconduct (especially authorship issues and fraudulent peer-review) took longer to retract than honest error, which was also observed in previous studies ^[22-24,30-33]^.

Just as commented by Alison McCook on the Retraction Watch blog ^[34]^, retractions due to fabricated peer reviews are becoming a trend. With the current global research output with 20,000 journals publishing more than 2 million articles per year and thousands of scientists publish a paper every five days ^[35-36]^, researchers worldwide are under great pressure to publish papers indexed by SCI for graduation, promotion, and acquisition of research funds. The need to publish has grown beyond the capacity of the scientists ^[37-38]^. Peer review aims to ensure that articles are “open and honest”, but it can be compromised when editors rely on authors’ recommended reviewers. This form of misconduct is particularly intense in China, but it would be a mistake to look at this as a Chinese or Asian problem. In this case, it is of paramount importance to understand that incentives really work, but the problem is that the perverse incentive systems in scientific publishing exists almost everywhere and may increase the risk of scientific misconduct.

We also found that unreliable data was one of the important reasons for the retraction, involving more than one fourth of the studies, and some SRs included retracted primary studies ^[39-41]^. SRs have the potential to provide sufficient sample size and generalizable population information to make more powerful evidence-based conclusions. Thus, great care must be used in the data analysis and determination of eligible studies. The methodology for conducting SRs has advanced enormously in the recent 30 years, authors should strengthen the study, and journals should refuse to publish SR not meeting rigorous standards ^[42-44]^. It is recommended to use the Preferred Reporting Items for Systematic Reviews and Meta-Analyses (PRISMA) statement and A Measurement Tool to Assess Systematic Reviews (AMSTAR) tool for the design, reporting and evaluation of SRs ^[45-47]^. More importantly, previous studies have shown that retracted literature continues to be cited as valid and legitimate work in many scientific disciplines, even after flagged as retracted ^[48]^. SRs are cornerstones of clinical practice guidelines in the medical field, and if retracted SRs are cited, the recommendations will be affected. Especially for SRs related to critical conditions, this will pose a great threat to medical care ^[49]^. Therefore, appropriate production, publication and citation of high-quality studies is essential for clinical decision-making and research.

In addition, our analyses suggested that COPE guidelines on retraction were not adhered to all respects. We found that not all retracted SRs had a correspondence notice or “retracted” label, and some retraction notices did not explicitly state the retraction reasons and who issued the retraction. In addition, we classified issuers into editor, author, published and journal as notice reported, but it maybe difficult to distinguish many of these, because the editor represents the journal that is owned by the publisher, therefore, these are basically the same entity. Publication of a clear and unambiguous retraction notice is an important part of the retraction, and the notice should be clearly identifiable, freely available, published promptly and linked to the original article that is retracted (which also should be clearly labeled as retracted) ^[50]^.

### Further suggestions

Regardless of the reasons for retraction, retracted SRs sometimes continue to be cited, so retracting such papers alone is not enough. It is important to recognize the dangers related to citing retracted literature and have clear guidance about how to address this issue. We propose the following interim recommendations:

Authors should strengthen and refine the learning of methodology, and strictly follow the relevant guidelines and PRISMA statement to improve the quality of conducting SRs from determining the clinical question until the reporting of the results. We recommend open registration of all SRs to reduce the risk for duplicate work ^[51]^. We emphasize the need to adhere to the principles of “5 dont’s of academic publishing”: ghostwriting, ghost submission, ghost revision, fake peer review, and falsified authorship. And it should be standard practice in all SRs to double-check the final list of included studies against the list of retracted articles ^[52]^. On the other hand, more detailed guidance on the inclusion/exclusion of retracted articles in SRs is needed, especially when the reason for retraction is not directly associated with a risk of inaccurate data, which are unlikely to affect the SR results ^[53]^.

For journal editors and publishers, the quality of peer review should be improved by shortening the period and make changes. It could be possible to do a bit more background check when selecting reviewers (even if first follow the recommendations), and probably combine with artificial intelligence. Experienced librarian, information specialist, or other search strategy experts can also be invited as methodological peer reviewers ^[54]^. Data should be strictly screened and checked before the manuscript is published to prevent publishing plagiarized studies, or studies with erroneous or falsified data, and most importantly, systematic and organized publication process manipulation should be altered and tackled ^[50]^. We recommend to follow or adopt a checklist (such as COPE guidelines) and templates to provide adequate information related to the retractions, so that readers can know who retracted the article and why the findings are considered unreliable. On the other hand, if an included study gets retracted after the SR was published, issuing a correction should be considered and giving the authors chance to update their SR.

For national and other academic institutions, the key to stemming the flow of poor-quality misleading research requires a unified and standardization criterion and a change in academic culture. First, national academic committees should strengthen the development of a credibility system, and put in place relevant laws and regulations. Second, for studies with serious academic misconduct (such as data falsification and faked peer review), an effective system of precautions and penalties should be implemented. National academic committees and research institutes might consider revoking research funding for such authors and demoting them. Third, academic institutions including but not limited to universities and hospitals should provide scientific ethics and academic integrity training to students, teachers, clinicians and other researchers, clarify the purpose of scientific research and reform unreasonable incentive mechanisms (for example, changing the way research success is measured and changing the way journal and article quality is measured, although in this study we used IF which maybe widely criticized).

For evidence users including health professionals and other stakeholders, especially guideline developers, methodological experts should always be included in the formulation of recommendations to rigorously review the cited SRs in terms of quality and make the decision whether to directly adopt, update or reproduce the review, and ensure the quality of evidence and that recommendations are not affected by the retracted publications ^[55-56]^. Clinicians, public health officials and other investigators can use AMSTAR tool to assess the methodological quality ^[47]^.

### Strengths and limitations

This study is to our knowledge the first to characterize the retraction status and reasons among non-Cochrane SRs in the field of medicine. We divided the risk of citation of retracted SRs into three categories according to the influence they can have on other studies. However, our study had also some limitations. First, we only searched the major international databases and websites, and did not search for example locally or regionally databases for research in other languages. Second, with so many SRs and rapid reviews to be published, especially in the recent setting of COVID-19, some of the studies may still become retracted so that the status and reasons may be changed.

## Conclusions

An increasing number of non-Cochrane SRs in the medical field are faced with the risk of retractions. China is the leading country of origin for retracted articles. The most common reasons for retraction are fraudulent peer-review and unreliable data. The average time from publication to retraction was 14 months, but some SRs were retracted even as much as nine years after publishing. Adherence to the COPE guidelines needs to be improved to provide adequate information and increase transparency of the retraction process, and authors should strengthen their knowledge in methodology.

## Supporting information

Supplementary Material for Search Strategy

## Data Availability

All data referred to in the manuscript was extracted from the included studies.

## Acknowledgments

We thank Janne Estill, Institute of Global Health of University of Geneva for providing guidance and comments for our review. We thank all the authors for their wonderful collaboration.

## Author contributions

Y.L.C. and Q.L.G. designed the paper and were consulted to revise the manuscript. S.Q.L. selected the literature, extracted data, carried out the statistical analysis, produced the tables and figures, and wrote the first edition of the paper. W.Z.J., Q.Z., R.Z.H., X.G. and S.E.H. helped selected the literature and extracted data. S.Y.Z. helped statistical analysis. M.Y.F. helped modify the first edition of the paper. All authors contributed to the review and approval of the final manuscript.

## Patient consent for publication

Not required.

