## Supplementary Material for Search Strategy for "An Overview of Retraction Status and Reasons of Non-Cochrane Systematic Reviews in Medicine"

**Supplementary Material I：Search Strategy**

**MEDLINE**

1. “Retraction of Publication as Topic” [Mesh]

2.“Retraction of Publication” [Publication Type]

3.“Retracted Publication” [Publication Type]

4. “Retract*” [Title/Abstract]

5.“Withdraw*” [Title/Abstract]

6. “Remov*” [Title/Abstract]

7. OR/ 1~6

8.“Systematic Reviews as Topic” [Mesh]

9. “Systematic Review” [Publication Type]

10."Meta-Analysis as Topic"[Mesh]

11. “meta-analys*” [Title/Abstract]

12.“metaanalys*” [Title/Abstract]

13.“systematic review*” [Title/Abstract]

14.“systematic literature review*” [Title/Abstract]

15.“systematic scoping review*” [Title/Abstract]

16.“systematic narrative review*” [Title/Abstract]

17.“systematic qualitative review*” [Title/Abstract]

18.“systematic evidence review*” [Title/Abstract]

19.“systematic quantitative review*” [Title/Abstract]

20.“systematic meta-review*” [Title/Abstract]

21.“systematic critical review*” [Title/Abstract]

22.“systematic mixed studies review*” [Title/Abstract]

23.“systematic mapping review*” [Title/Abstract]

24.“systematic cochrane review*” [Title/Abstract]

25.“systematic integrative review*” [Title/Abstract]

26. OR/ 8~25

27. 7 AND 26

**Embase**

1. 'retraction'/exp
2. 'retraction notice'/exp
3. 'retract*':ti
4. 'withdraw*':ti
5. 'remov*':ti
6. OR/ 1~5
7. 'meta analysis (topic)'/exp
8. 'meta analysis'/exp
9. 'systematic review'/exp
10. 'systematic review (topic)'/exp
11. 'meta-analys*':ti
12. 'meta analys*':ti
13. 'systematic review*':ti
14. 'systematic literature review*':ti
15. 'systematic scoping review*':ti
16. 'systematic narrative review*':ti
17. 'systematic qualitative review*':ti
18. 'systematic evidence review*':ti
19. 'systematic quantitative review*':ti
20. 'systematic meta-review*':ti
21. 'systematic critical review*':ti
22. 'systematic mixed studies review*':ti
23. 'systematic mapping review*':ti
24. 'systematic cochrane review*':ti
25. 'systematic integrative review*':ti
26. OR/ 7-25
27. 6 AND 26
28. 27 AND [medline]/lim
29. 27 NOT 28
